# Consistency of performance of adverse outcome prediction models for hospitalized COVID-19 patients

**DOI:** 10.1101/2021.02.25.21252410

**Authors:** Victor M. Castro, Thomas H. McCoy, Roy H. Perlis

**Author notes:** **Correspondence:** Roy H. Perlis, Massachusetts General Hospital, 185 Cambridge Street, 6th Floor, Boston, MA 02114.

## Abstract

**Background:** We previously reported and validated a risk prediction tool based on COVID-19 hospitalizations prior to June 2020. Here, we report performance of that model on subsequent data from 6 hospitals and among individual patient subgroups.

**Method:** We included individuals age 18 or older hospitalized at one of 2 academic medical centers and 4 community hospitals from 6/7/2020 through 1/22/2021 with positive PCR test for severe acute respiratory syndrome coronavirus 2 (SARS-CoV-2) within 5 days of admission. Coefficients from our previously reported least absolute shrinkage and selection operator (Lasso) risk models were applied to estimate probability of a mortality, as well as a composite severe illness outcome, including admission to the ICU, mechanical ventilation or mortality.

**Results:** Overall model performance for mortality included AUC of 0.83 (95% CI:0.80-0.87) for mortality, with a PPV 0.55 and NPV of 0.95 when using a cutoff corresponding to the highest 20% of predicted risk derived in the training set. For all adverse outcomes, AUC was 0.79 (95% CI:0.75-0.81) and PPV 0.48 and NPV 0.98 in the top 20% risk group. Model discrimination was generally similar between genders and race/ethnicity groups, but markedly poorer for younger age groups.

**Conclusion:** Although the population of individuals hospitalized for COVID-19 has shifted and outcomes have improved overall, prediction models derived earlier in the pandemic may maintain utility.

## Introduction

The challenge of managing limited resources during COVID-19 has sparked efforts to stratify risk among hospitalized patients^1^. Few risk models have been validated or investigated for potential bias,^2^ even though COVID-19 inpatient populations, treatments, and outcomes have changed over time. We previously reported and validated a risk prediction tool based on COVID-19 hospitalizations prior to June 2020^3^. Here, we report performance of that model on subsequent data from 6 hospitals and among individual patient subgroups.

## Method

We included individuals age 18 or older hospitalized at one of 2 academic medical centers and 4 community hospitals from 6/7/2020 through 1/22/2021 with positive PCR test for severe acute respiratory syndrome coronavirus 2 (SARS-CoV-2) within 5 days of admission, excluding those with an outcome on the day of hospitalization. Features of hospital course were extracted from the Mass General Brigham Data Registry^4^ and the Enterprise Data Warehouse, including laboratory values and high/low flags. Charlson comorbidity index was calculated using coded ICD-10 diagnostic codes^5^. The study protocol was approved by the Mass General Brigham Human Research Committee, waiving informed consent as detailed by 45 CFR 46.116. STROBE reporting guidelines for cohort studies were applied. Patients were followed from admission to hospital discharge or death, with follow-up censored at discharge. Primary outcomes of interest were 1) a composite severe illness outcome, including admission to the ICU, mechanical ventilation or mortality; and 2) mortality. Coefficients from our previously reported least absolute shrinkage and selection operator (Lasso) risk models were applied to estimate probability of each outcome, without recalibration. We applied median imputation of missing data. We characterized model performance with standard metrics of discrimination and calibration. All analyses utilized R v4.

## Results

Features of the new cohort are summarized in Table 1 and compared to those of the previously-reported cohort in which the predictive model was trained. For the 2,892 individuals, mean age was 63.0 (SD=19.1); they were 50.5% female, 23.3% Hispanic, and 11.9% Black. Mean length of hospital stay was 6.2 days (SD=5.3); 4.4% required ICU stay and 2.4% mechanical ventilation, while 5.8% died prior to discharge. Overall model performance for mortality included AUC of 0.83 (95% CI:0.80-0.87) for mortality, with a PPV 0.55 and NPV of 0.95 when using a cutoff corresponding to the highest 20% of predicted risk derived in the training set. For all adverse outcomes, AUC was 0.79 (95% CI:0.75-0.81) and PPV 0.48 and NPV 0.98 in the top 20% risk group. Among subgroups (Table 2), model discrimination was generally similar between genders and race/ethnicity groups, but markedly poorer for younger age groups.

**Table 1.**
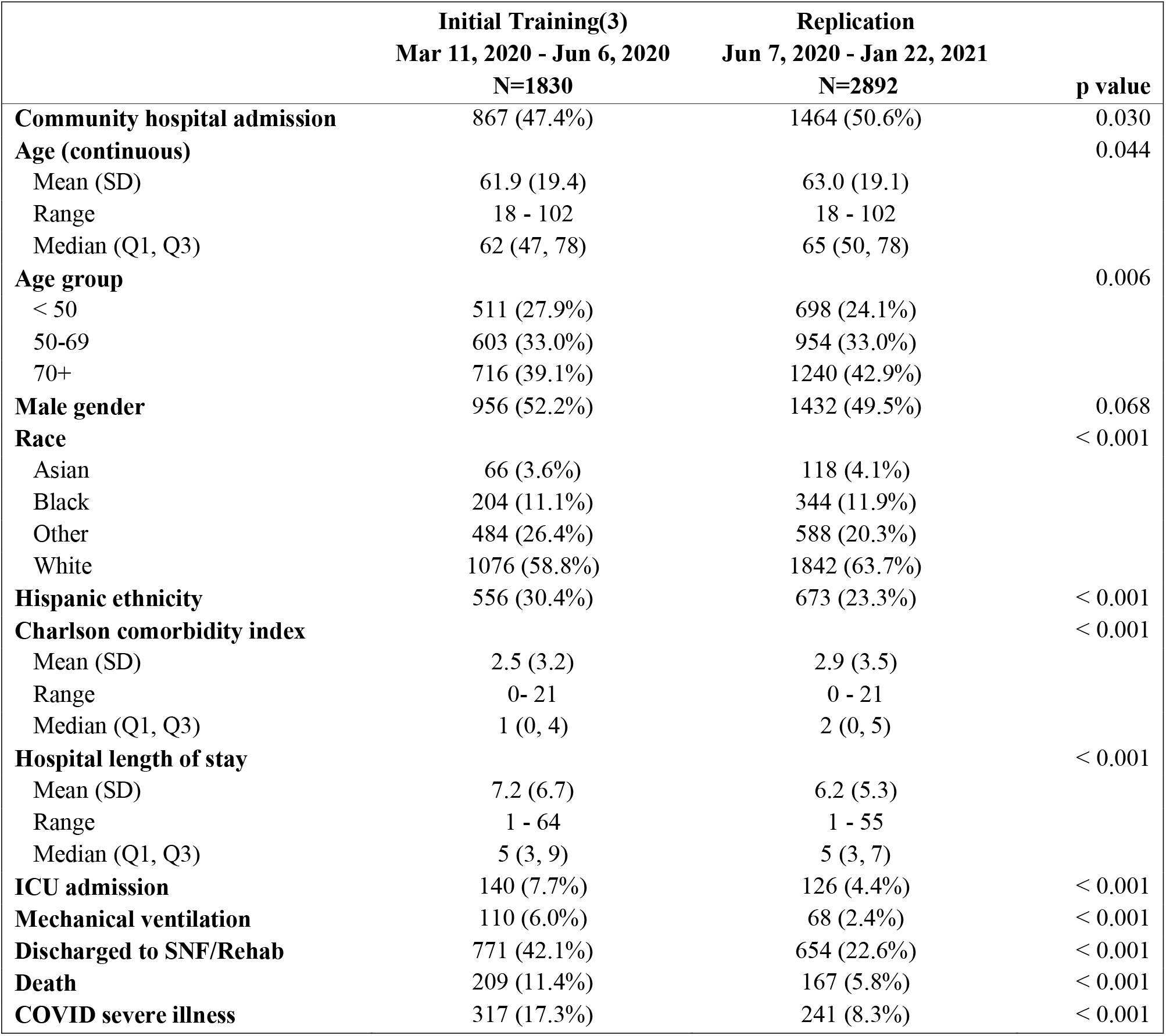
Sociodemographic and clinical feature comparison between the initial model training COVID admissions and the subsequent admissions used to evaluate the model. The training dataset consisted of the initial surge of COVID cases in Eastern Massachusetts whereas the replication cohort includes the summer nadir and second wave in Fall 2020.

|  | **Initial Training(3)**<br>**Mar 11, 2020 - Jun 6, 2020**<br>**N=1830** | **Replication**<br>**Jun 7, 2020 - Jan 22, 2021**<br>**N=2892** | **p value** |
| --- | --- | --- | --- |
| **Community hospital admission** | 867 (47.4%) | 1464 (50.6%) | 0.030 |
| **Age (continuous)** |  |  | 0.044 |
| Mean (SD) | 61.9 (19.4) | 63.0 (19.1) |  |
| Range | 18 - 102 | 18 - 102 |  |
| Median (Q1, Q3) | 62 (47, 78) | 65 (50, 78) |  |
| **Age group** |  |  | 0.006 |
| < 50 | 511 (27.9%) | 698 (24.1%) |  |
| 50-69 | 603 (33.0%) | 954 (33.0%) |  |
| 70+ | 716 (39.1%) | 1240 (42.9%) |  |
| **Male gender** | 956 (52.2%) | 1432 (49.5%) | 0.068 |
| **Race** |  |  | < 0.001 |
| Asian | 66 (3.6%) | 118 (4.1%) |  |
| Black | 204 (11.1%) | 344 (11.9%) |  |
| Other | 484 (26.4%) | 588 (20.3%) |  |
| White | 1076 (58.8%) | 1842 (63.7%) |  |
| **Hispanic ethnicity** | 556 (30.4%) | 673 (23.3%) | < 0.001 |
| **Charlson comorbidity index** |  |  | < 0.001 |
| Mean (SD) | 2.5 (3.2) | 2.9 (3.5) |  |
| Range | 0- 21 | 0 - 21 |  |
| Median (Q1, Q3) | 1 (0, 4) | 2 (0, 5) |  |
| **Hospital length of stay** |  |  | < 0.001 |
| Mean (SD) | 7.2 (6.7) | 6.2 (5.3) |  |
| Range | 1 - 64 | 1 - 55 |  |
| Median (Q1, Q3) | 5 (3, 9) | 5 (3, 7) |  |
| **ICU admission** | 140 (7.7%) | 126 (4.4%) | < 0.001 |
| **Mechanical ventilation** | 110 (6.0%) | 68 (2.4%) | < 0.001 |
| **Discharged to SNF/Rehab** | 771 (42.1%) | 654 (22.6%) | < 0.001 |
| **Death** | 209 (11.4%) | 167 (5.8%) | < 0.001 |
| **COVID severe illness** | 317 (17.3%) | 241 (8.3%) | < 0.001 |

**Table 2A.** Discrimination and calibration metrics of the COVID severity prediction model by subgroup

| Subgroup | Admissions | COVID<br>severe illness | ROC AUC<br>(95% CI) | PR AUC | Specificity | Sensitivity | PPV | NPV |
| --- | --- | --- | --- | --- | --- | --- | --- | --- |
| **Academic medical centers** | 1,428 | 106 | 0.749 (0.696-0.802) | 0.286 | 1.00 | 0.05 | 0.62 | 0.93 |
| **Community hospitals** | 1,464 | 135 | 0.804 (0.768-0.839) | 0.273 | 1.00 | 0.02 | 0.33 | 0.91 |
| **Female** | 1,460 | 91 | 0.769 (0.717-0.821) | 0.211 | 1.00 | 0.02 | 0.29 | 0.94 |
| **Male** | 1,432 | 150 | 0.775 (0.736-0.815) | 0.331 | 1.00 | 0.04 | 0.60 | 0.90 |
| **< 50** | 698 | 17 | 0.638 (0.481-0.794) | 0.083 | 1.00 | 0.00 | - | 0.98 |
| **50-69** | 954 | 58 | 0.691 (0.623-0.759) | 0.127 | 1.00 | 0.00 | 0.00 | 0.94 |
| **70+** | 1,240 | 166 | 0.768 (0.730-0.805) | 0.346 | 0.99 | 0.05 | 0.50 | 0.87 |
| **Asian** | 118 | 10 | 0.865 (0.747-0.983) | 0.476 | 1.00 | 0.00 | - | 0.92 |
| **Black** | 344 | 20 | 0.740 (0.627-0.853) | 0.158 | 1.00 | 0.00 | 0.00 | 0.94 |
| **Other** | 588 | 30 | 0.766 (0.677-0.856) | 0.235 | 0.99 | 0.13 | 0.57 | 0.96 |
| **White** | 1,842 | 181 | 0.776 (0.74-0.812) | 0.294 | 1.00 | 0.02 | 0.44 | 0.90 |
| **Hispanic** | 673 | 30 | 0.736 (0.629-0.843) | 0.135 | 0.99 | 0.07 | 0.29 | 0.96 |
| **Not Hispanic** | 2,219 | 211 | 0.779 (0.747-0.812) | 0.311 | 1.00 | 0.03 | 0.60 | 0.91 |

**Table 2B.** Discrimination and calibration metrics of the COVID mortality prediction model by subgroup

| Subgroup | Admissions | Died in<br>Hospital | ROC AUC<br>(95% CI) | PR AUC | Specificity | Sensitivity | PPV | NPV |
| --- | --- | --- | --- | --- | --- | --- | --- | --- |
| **Academic medical centers** | 1,428 | 70 | 0.800 (0.740-0.860) | 0.290 | 1.00 | 0.07 | 0.63 | 0.95 |
| **Community hospitals** | 1,464 | 97 | 0.859 (0.824-0.893) | 0.316 | 0.99 | 0.07 | 0.50 | 0.94 |
| **Female** | 1,460 | 60 | 0.822 (0.762-0.882) | 0.201 | 0.99 | 0.07 | 0.33 | 0.96 |
| **Male** | 1,432 | 107 | 0.835 (0.795-0.875) | 0.381 | 1.00 | 0.08 | 0.80 | 0.93 |
| **< 50** | 698 | 7 | 0.556 (0.312-0.800) | 0.032 | 1.00 | 0.00 | - | 0.99 |
| **50-69** | 954 | 27 | 0.702 (0.605-0.799) | 0.064 | 1.00 | 0.00 | 0.00 | 0.97 |
| **70+** | 1,240 | 133 | 0.812 (0.776-0.849) | 0.370 | 0.99 | 0.09 | 0.57 | 0.90 |
| **Asian** | 118 | 6 | 0.899 (0.803-0.995) | 0.377 | 1.00 | 0.17 | 1.00 | 0.96 |
| **Black** | 344 | 11 | 0.901 (0.842-0.961) | 0.218 | 1.00 | 0.00 | 0.00 | 0.97 |
| **Other** | 588 | 17 | 0.839 (0.725-0.953) | 0.292 | 0.99 | 0.18 | 0.37 | 0.98 |
| **White** | 1,842 | 133 | 0.814 (0.774-0.854) | 0.328 | 1.00 | 0.06 | 0.67 | 0.93 |
| **Hispanic** | 673 | 16 | 0.751 (0.596-0.905) | 0.170 | 0.99 | 0.06 | 0.17 | 0.98 |
| **Not Hispanic** | 2,219 | 151 | 0.834 (0.801-0.867) | 0.336 | 1.00 | 0.07 | 0.69 | 0.94 |

ROC AUC: Area under the receiver operating characteristic curve, CI: Confidence interval; PR AUC: Area under the precision recall curve, PPV: Positive predictive value, NPV: Negative predictive value. Specificity, Sensitivity, PPV and NPV are reported for the top 20% of risk score defined in the training set.

## Discussion

Applying a previously-validated model to 2,892 new COVID-19 admissions in the same 6 hospitals, we find that model performance decreased modestly from the initial validation study which reported AUC’s of 0.81 for severe illness and 0.85 for mortality^3^. Discrimination is generally similar across subgroups, with the notable exception of younger age groups, where performance is poorer.

Our results highlight the observation that the population of individuals hospitalized for COVID-19 has shifted and outcomes have improved overall, but suggest that prediction models derived earlier in the pandemic may maintain utility. The extent to which this finding of robustness over time in a rapidly evolving disease generalizes to other predictive models requires investigation. Our results also illustrate the importance of investigating risk stratification models across patient subgroups, as a step toward ensuring that particular groups of patients are not adversely impacted by application of such tools particularly in settings of potential resource constraints. Models that explicitly address the heterogeneity in severe COVID-19 across age groups are candidates for further study.

## Data Availability

The hospital institutional review board has not approved data release outside of the institution.

## Acknowledgements

This study was supported by the National Institute of Mental Health (R01MH116270; Dr. Perlis). The sponsors did not contribute to design and conduct of the study; collection, management, analysis, and interpretation of the data; preparation, review, or approval of the manuscript; and decision to submit the manuscript for publication. Dr. Perlis had full access to all the data in the study and takes responsibility for the integrity of the data and the accuracy of the data analysis. He is an associate editor for *JAMA Network Open*.

